# Large language models’ interpretation homogeneity and text Analysis: Evaluating the utility of the global flu view platform for Influenza surveillance

**DOI:** 10.1101/2025.01.12.25320440

**Authors:** Joy Luzingu, Iman Hakim, Onicio Leal Neto

## Abstract

The advent of Large Language Models (LLMs) has transformed natural language processing and offers new possibilities for analyzing qualitative data in public health research. This study evaluated the utility of Global Flu View (GFV), a participatory surveillance platform for influenza-like illness, using multiple LLMs to analyze stakeholder perceptions. We conducted in-depth interviews with 10 participants comprising GFV partners, advisory group members, and public health researchers across six countries. Interview transcripts were analyzed using four LLMs (ChatGPT, Claude AI, Perplexity, and Gemini) to perform sentiment analysis, with scores ranging from -1 (negative) to +1 (positive). Word pair networks were generated using the Louvain clustering method to identify thematic patterns in stakeholder responses. The analysis revealed consistently positive sentiments toward GFV across all stakeholder groups, with sentiment scores ranging from 0.4 to 0.8. The Friedman test showed that ChatGPT and Gemini produced higher sentiment scores (average rank = 3.5 each) compared to Claude AI (average rank ≈ 1.33) and Perplexity (average rank ≈ 1.67). Word pair networks demonstrated that participants conceptualize GFV as a data-centric tool integrated within broader public health systems, with strong connections between terms related to surveillance, global collaboration, and public health decision-making. While stakeholders expressed optimism about GFV’s potential to enhance global influenza surveillance, they also identified areas for improvement, particularly regarding data sharing and regional implementation. This study represents the first evaluation of GFV using LLMs for sentiment analysis and demonstrates the potential of AI-assisted qualitative analysis in public health research. The findings suggest that GFV is perceived as a valuable tool for global health surveillance, while also highlighting opportunities for platform enhancement and methodological considerations for future AI-assisted qualitative research in public health.

**Author summary:** In our study, we explored how public health experts view and value a new global health surveillance platform called Global Flu View, which collects data about flu-like illnesses from multiple countries across four continents. We were particularly interested in understanding whether this platform is helping to track and monitor flu-like illnesses worldwide. To do this, we interviewed 10 experts from different countries and used artificial intelligence tools to analyze their responses. This approach allowed us to systematically evaluate their perceptions and sentiment towards the platform. We found that experts generally view the platform positively and see it as a valuable tool for global health monitoring. However, they also pointed out areas where it could be improved, such as making data sharing between countries easier. Our work is important because it helps us understand how new digital tools can support global health surveillance, and it demonstrates how artificial intelligence can help researchers analyze interview data. The insights we gained can help make these kinds of health monitoring platforms more effective and useful for tracking disease outbreaks across the world.

## 1. Introduction

Public health disease surveillance will be drastically changed because of the advent of Large Language Models (LLMs). In recent years this has significantly transformed the landscape of natural language processing (NLP), a field of artificial intelligence (AI) whose purpose is to enable computers to ‘understand’ and generate human-readable text [1, 2]. On November 30, 2022, a groundbreaking advancement in AI emerged with the introduction of ChatGPT by OpenAI Inc. This generative pretrained transformer (GPT) technology garnered remarkable attention, reaching 100 million users within just three months, a record-breaking achievement for any internet application [3]. Driven by the popularity of ChatGPT, LLMs are gaining attention as tools for advancing research in health sciences [4, 5]. For example, Meditron, an open-source domain adapted medical LLM, demonstrates the potential of LLMs in medicine and public health by leveraging domain-specific data (clinical guidelines, paper abstracts, medical papers, and replay dataset) to address healthcare challenges [6]. Generative AI models may align particularly well with the need of qualitative research. These models’ capacity to rapidly process and synthetize large volumes of text, combined with their ability to distill diverse inputs into unified themes, offers promising possibilities for researchers seeking to expand and enrich their analytical approaches [7]. Furthermore, their content-sensitive reasoning capabilities mirror human cognitive patterns, enhancing their utility in qualitative analysis [8].They can help qualitative researchers in many ways: hypothesis generation, coding and thematic analysis, literature review and synthesis, writing, and data analysis [7]. ChatGPT has demonstrated remarkable performance in a set of tasks, comprising language translation, text completion and sentiment analysis, with its ability to generate coherent and appropriated answers to text inputs, making it a valuable tool for a large range of applications, including public health [9]. Research has suggested that AI could be a promising tool for qualitative research [10, 11]. Thus, these LLMs can be used to extract sentiment from public health experts regarding various topics, including the utility of tools used in prevention against infectious diseases and enhancing public health intelligence (PHI) which is defined as “a core public health function responsible for identifying, collecting, connecting, synthetizing, analyzing, assessing, interpreting and generating a wide range of information for actionable insights and disseminating these for informed and effective decision-making to protect and improve the health of the population”[12].

Influenza-Like Illness (ILI) continues to pose a significant global health challenge, causing up to 650,000 respiratory-related deaths annually and substantial economic burdens on healthcare systems worldwide [13]. It represents a major global health issue that requires coordinated surveillance efforts across various geographic regions to rapidly detect potential outbreaks and inform necessary response strategies [14]. Timely detection and surveillance are critical for mitigating its impacts, particularly in a rapidly interconnected word where influenza outbreaks can cross national boundaries within days. Traditional surveillance systems, which rely on clinical reports and laboratory data, often face limitations in coverage, timeliness, and data details. To address these challenges, participatory surveillance platforms have emerged as innovative tools that harness the power of digital technology and community engagement to collect real-time data on ILI symptoms from the general population [15, 16]. The advent of participatory surveillance platforms, a key topic in digital epidemiology [17], has revolutionized the collection and use of public health data. These platforms enable individuals to report symptoms and health behaviors in real time, complementing traditional systems with richer and more immediate datasets. Notable examples include Flutracking in Australia, InfluenzaNet across Europe and Outbreak Near Me (formerly known as Flu Near you) in North America [18].

Historically, one of the first use of crowdsourcing for public health surveillance was started in 2003 in the Netherlands during the 2003-2004 influenza season. In fact, in September 2003, an internet-based Great Influenza Survey (GIS) was initiated in the Netherlands and the dutch-speaking part of Belgium with the objective of rendering scientific information accessible to the general public and ignite students’ enthusiasm for science. Participants were invited through weekly emails to provide their demographic information and answer questions about ILI symptoms they might have experienced. In return, the GIS website regularly provided up-to-date feedback information on how the country was evolving during the influenza season. As a result, comparing the ILI rates between GIS and the Dutch sentinel practice network operated by general practitioners, a strong similarity between the two seasonal courses of ILI measured by the two systems was found, with higher rate of ILI in GIS but the curves having almost a similar pattern[19]. A couple of years later, the 2009 H1N1 pandemic, the first influenza pandemic in more than 4 decades [20] occurred, and exposed vulnerabilities in traditional surveillance systems, including delays in production of estimates of the total disease burden associated with the virus during the annual seasonal influenza epidemics [21]. This shortcoming underscored the need for tools that leverage digital technologies to enhance timeliness, geographic reach, and inclusivity.

Among ILI participatory surveillance systems, the Global Flu View (GFV) platform stands out as a transformative tool for global influenza surveillance. It represents a breakthrough in participatory surveillance integration, functioning as a system orchestrator that harmonizes data from multiple participatory surveillance organizations. Through collaborative development, participating organizations have established a shared Application Programming Interface (API) that facilitates data exchange among systems [18]. This integration has resulted in the first-of-its-kind global surveillance network, combining ILI data from more than 10 countries and Hong Kong across four continents [22], and generating thousands of weekly reports accessible through interactive visualization tools including graphs, maps, and customizable filters on the website www.globalfluview.org [18]. A key feature of GFV is the Codelab, a space reserved for program partners (experts, data scientists and public health practitioners), allowing them to share and improve their codes, scripts and methods used on participatory disease surveillance data [22]. GFV is an integrated platform that not only can detect flu-activity at global level early but also inform public health decision-making with actionable data. It was first launched as a pilot in 2019 and became publicly available globally in 2022.

Despite GFV’s innovative approach to global influenza surveillance, its utility and perceived effectiveness in supporting global influenza surveillance and informing public health decision-making have yet to be comprehensively evaluated. The objective of this study is to assess the utility of GFV and gather insights into its perceived effectiveness from the perspective of its partners, advisory group members, and public health researchers, using LLMs. By examining their experiences and viewpoints, this study aims to inform future enhancements to GFV and contribute to the broader understanding of the role of participatory surveillance in global health initiatives.

## 2. Material and Methods

### 2.1. Study design

This study used a mixed-methods approach to explore perceptions and experiences of stakeholders and public health experts with the GFV platform using four LLMs, namely ChatGPT (Open AI)[23], Claude AI (Anthropic)[24], Perplexity (Perplexity AI)[25] and Gemini (Google)[26].

### 2.2. Study context

This study was conducted between May and October 2024 at the University of Arizona, in Tucson in the framework of the GFV Spark program utility goal [27]. This program aimed at assessing the perceived utility of GFV from key stakeholders, exploring possibilities of enhancements in the platform, and allowing the selected student to interview public health officials, researchers, and decision-makers, and interact with existing program partners to identify strengths, weaknesses, and opportunities of improvement.

### 2.3. Study participants

Seventeen stakeholders were recruited by email to participate in the study based on their previous work and knowledge of GFV, as well as expertise on participatory surveillance, allowing them to provide reliable responses to questions. Seven were GFV program partners representatives, three were advisory group members, and seven were public health researchers. From those seventeen, ten participants accepted to be enrolled and were interviewed. They included five GFV partners representatives, two advisory group members, and three public health researchers, scattered across multiple countries, namely the US, Australia, Thailand, Hong Kong, England, and Italy (Fig.1).

**Figure 1.**
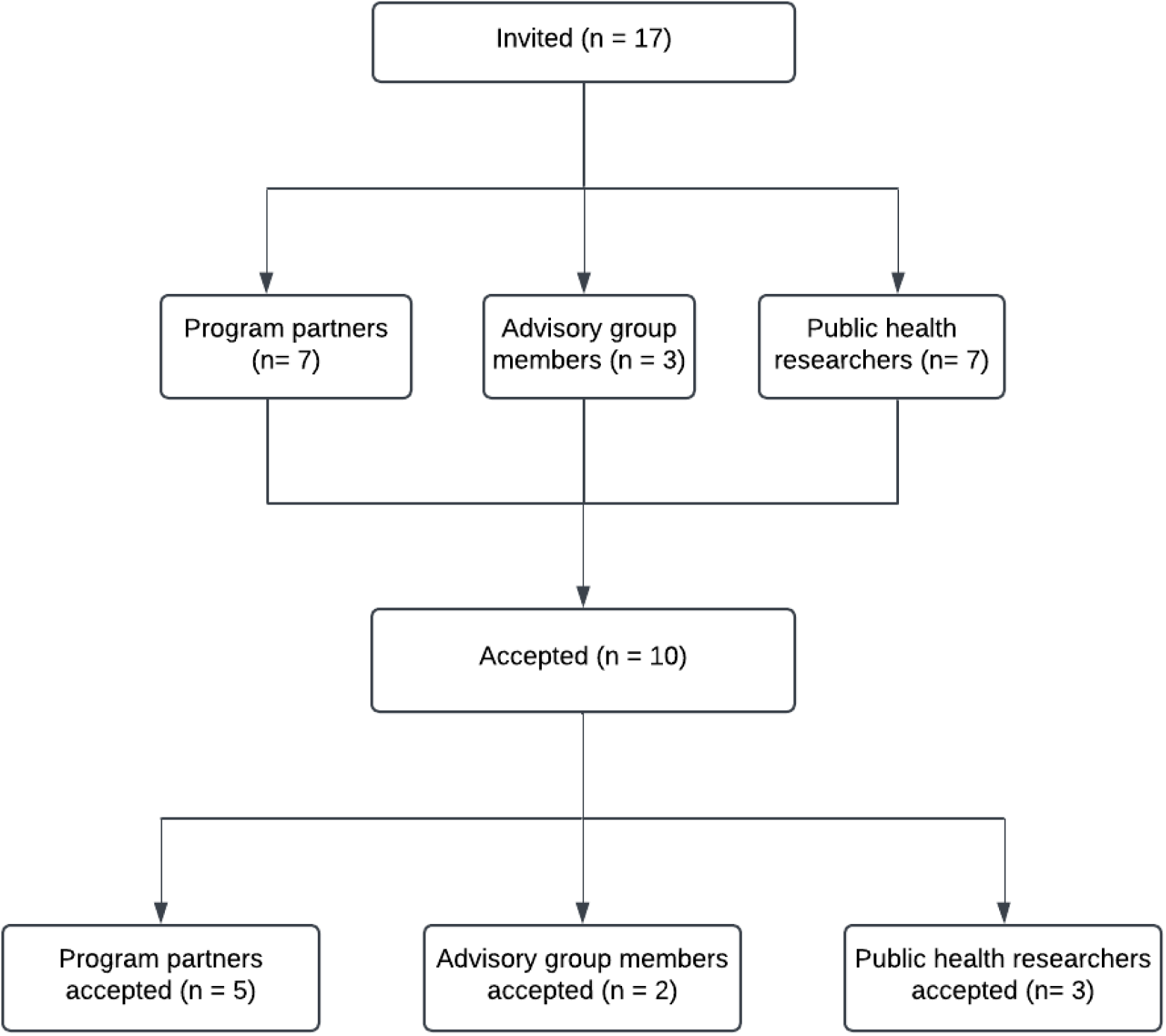
Participant flow diagram

### 2.4. Study procedures

Data were collected through in-depth interviews using three different interview guides tailored to the three groups of study participants (GFV partners representatives, advisory group members, and public health researchers). The interview guides for GFV partners representatives (Supplementary material 1) and public health researchers (Supplementary material 2) comprised 10 questions for each, and that of advisory group members included 8 questions (Supplementary material 3). Questions focused on key areas of interest: (1) participant’s perception of GFV as a system orchestrator and data hub, (2) their experience using the platform, (3) challenges encountered in utilizing GFV tool and accessing ILI data, and (4) future directions for the platform. For the term “system orchestrator”, we defined as a *“platform that automates and handles flow of data across multiple systems with a data ecosystem”*. And the definition used for “data hub” was a *“centralized platform that acts as a central point for managing and integrating data from various sources within an organization*”. Individual interviews were conducted via Zoom, version 6.2.10 (43047) (Zoom Video Communications Inc, San Jose, CA, USA), a collaborative, cloud-based videoconferencing platform that offers features such as online meetings, chat, video conference and audio recordings, and meeting transcripts [28], with each session lasting approximately 30 minutes. All interviews were recorded and automatically transcribed through Zoom’s built-in transcription feature, and the resulting transcripts saved as text (.txt) for analysis.

### 2.5. Data wrangling

Text files of zoom-generated transcripts were pre-processed using an online spreadsheet tool to create three datasets based on the three participants groups. Each dataset comprised question numbers, respondents’ initials, respondents’ answers, and their gender. Time code and interviewer questions were removed. Finally, responses only were saved into three text (.txt) datasets based on participant type: GFV partners, advisory group members, and public health researchers.

### 2.6. Data analysis

#### 2.6.1. Descriptive analysis

Using Exploratory software, version 10.6 (Exploratory, Inc., Redwood City, CA, USA) [29], text data from the three datasets were analyzed using the library Quanteda [30] for a word-pair networks based on the Louvain clustering method. This method is a community detection algorithm used to identify groups of closely connected nodes within a large network by optimizing a quality function called “modularity”. Essentially it finds clusters where nodes within a group are more densely connected to each other than nodes in other groups. It works by iteratively assigning nodes to communities based on local optimization, then aggregating these communities into new nodes and repeating the process until no further improvement in modularity can be achieved [31]. Stop-word libraries were developed for the 3 datasets (Supplementary materials 4,5, and 6) to refine analyses and remove words that were not meaningful in the context of the interviews. Punctuations and numbers were also removed. The number of words for each word pairs network was set at 50.

#### 2.6.2. Prompt engineering and Sentiment analysis

Four generative AI platforms were employed to conduct sentiment analysis (SA) from participants responses, including ChatGPT (model GPT-4), Claude AI (model Claude 3.5 Sonnet Opus), Perplexity (model Claude 3.5 Sonnet), and Gemini (model Gemini 1.5 Pro). The datasets containing participants’ responses were uploaded to these platforms for analysis. Studies have emphasized the importance of crafting high-quality prompts for LLMs, as prompt that seem to convey similar instructions can result in notably different outcomes [32]. In this study, we used the same optimized prompt across the four platforms to generate sentiment score from each dataset, stating “*Please, on a spectrum ranging from -1 being negative to +1 being positive, how would you rate the sentiment from the interview answers in the attached dataset?*”. However, it is important to mention that this prompt was just an exploratory approach.

We used the Friedman test to compare results across multiple conditions while accounting for potential block effects. The Friedman test is a nonparametric alternative to repeated-measures ANOVA that ranks observations within each block (in our case, each block was defined by a particular grouping factor) and sums (or averages) those ranks for each treatment (e.g., each large language model). The Friedman test avoids assumptions of normality and equal variances due to its intended ranking instead of raw data. For each treatment, we computed an average rank by aggregating its rank values across all blocks. Under the null hypothesis that all treatments perform identically, the average ranks should be roughly equal. A statistically significant Friedman test statistic indicates that at least one treatment’s median response or distribution differs meaningfully from the others[33]. We ran the test in R, using the libraries ‘ggplot2’, ‘dplyr’and ‘reshape2’.

## Ethical considerations

The University of Arizona’s IRB deemed the study not to be research involving human subjects as defined by Department of Health and Human Services and Food and Drug Administration regulations and determined that IRB review was exempt (IRB ID: STUDY00004947). Before each interview, written informed consent was obtained from participants voluntarily. Each participant was informed of the study’s objectives, methods, benefits, and possible risks during the interview session. We maintained confidentiality of participants information throughout the study period. They were allowed to stop the interview at any time. However, all participants were engaged in the full interview and agreed to be Zoom-recorded for transcription purposes.

## 3. Results

### 3.1. Word pair networks

The co-occurrence networks in figures 3, 4, and 5 reveal how participants conceptualize GFV through their language choices. With ‘Data’ as the consistent central node across all word pair networks, participants demonstrate that they perceive GFV as a data-centric tool, while simultaneously they connect it to broader public health practices and systems. The strong co-occurrence patterns between terms like ‘surveillance’, ‘participatory’, ‘global’, and ‘health’ suggest participants view GFV not in isolation, but as an integral part of the larger public health monitoring ecosystem. The frequent connections to ‘countries’, ‘systems’, and ‘stakeholders’ indicate that participants particularly value GFV’s role in facilitating international collaboration and bridging geographic and institutional boundaries. Additionally, the semantic clustering of disease-related terms (’flu’, ‘respiratory’, ‘illness’) with implementation concepts (’system’, ‘platform’, ‘tool’) reflects how participants understand the translation of data into actionable public health insights.

**Figure 2.**
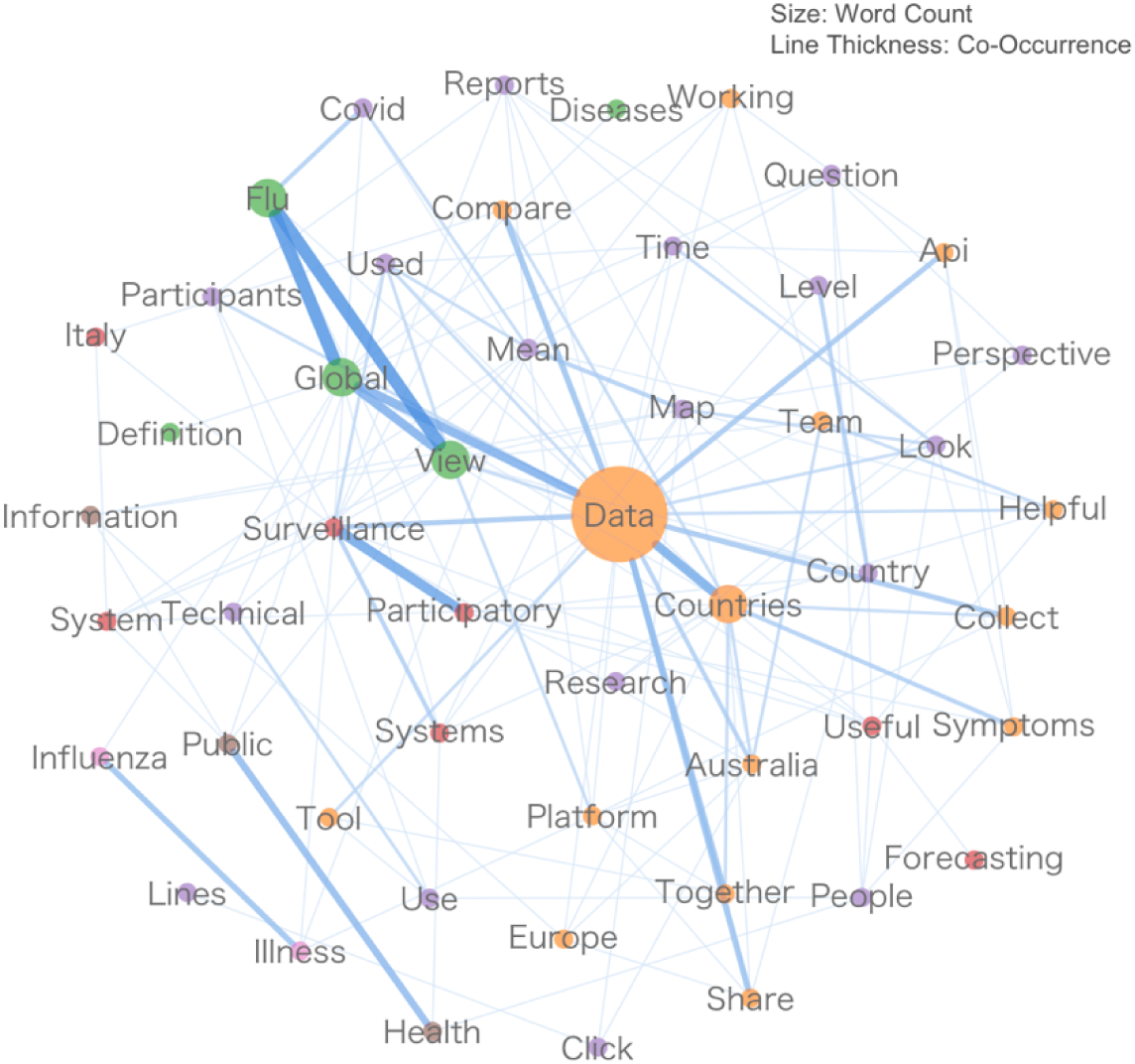
Word pair network from GFV partners

**Figure 3.**
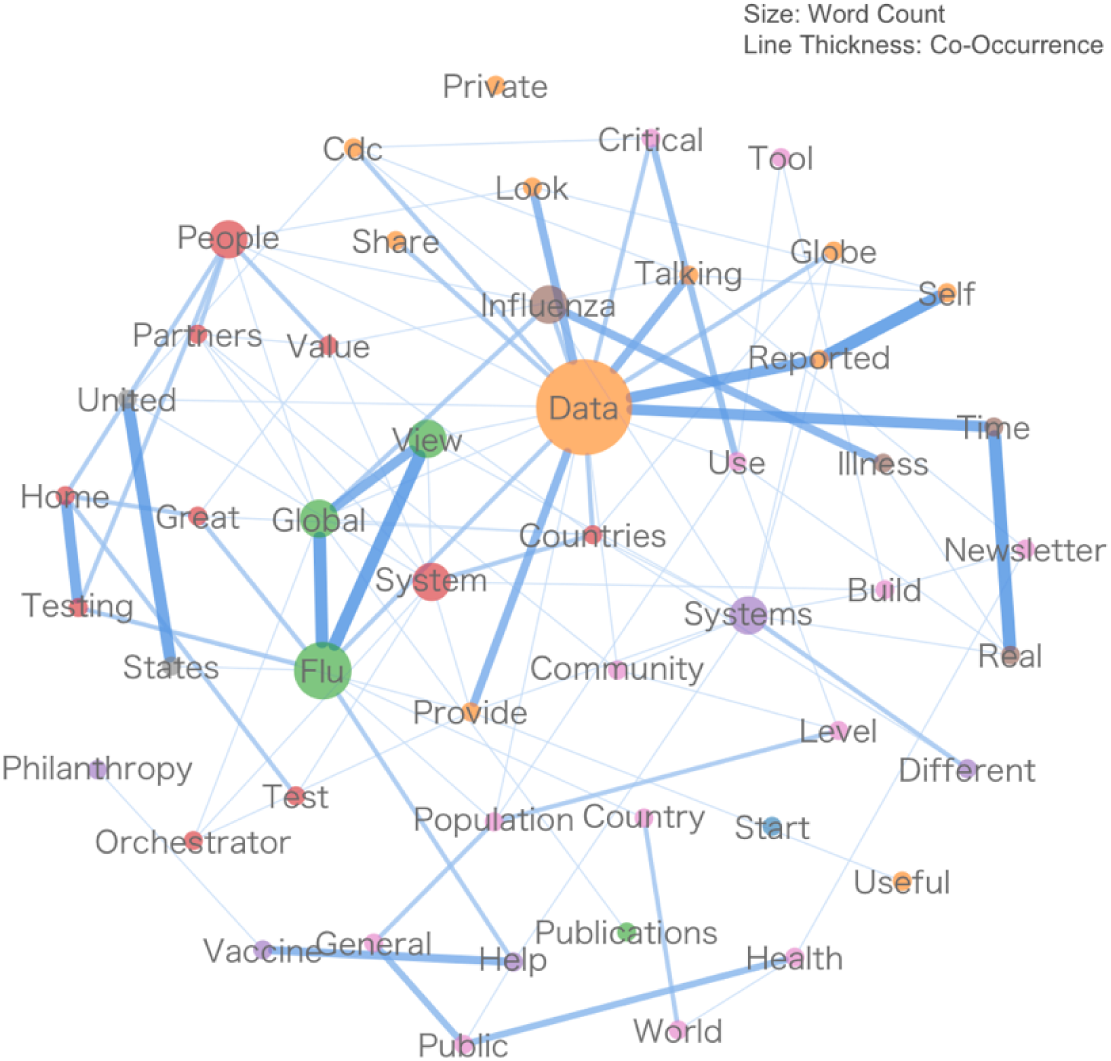
Word pair network from advisory group members

**Figure 4.**
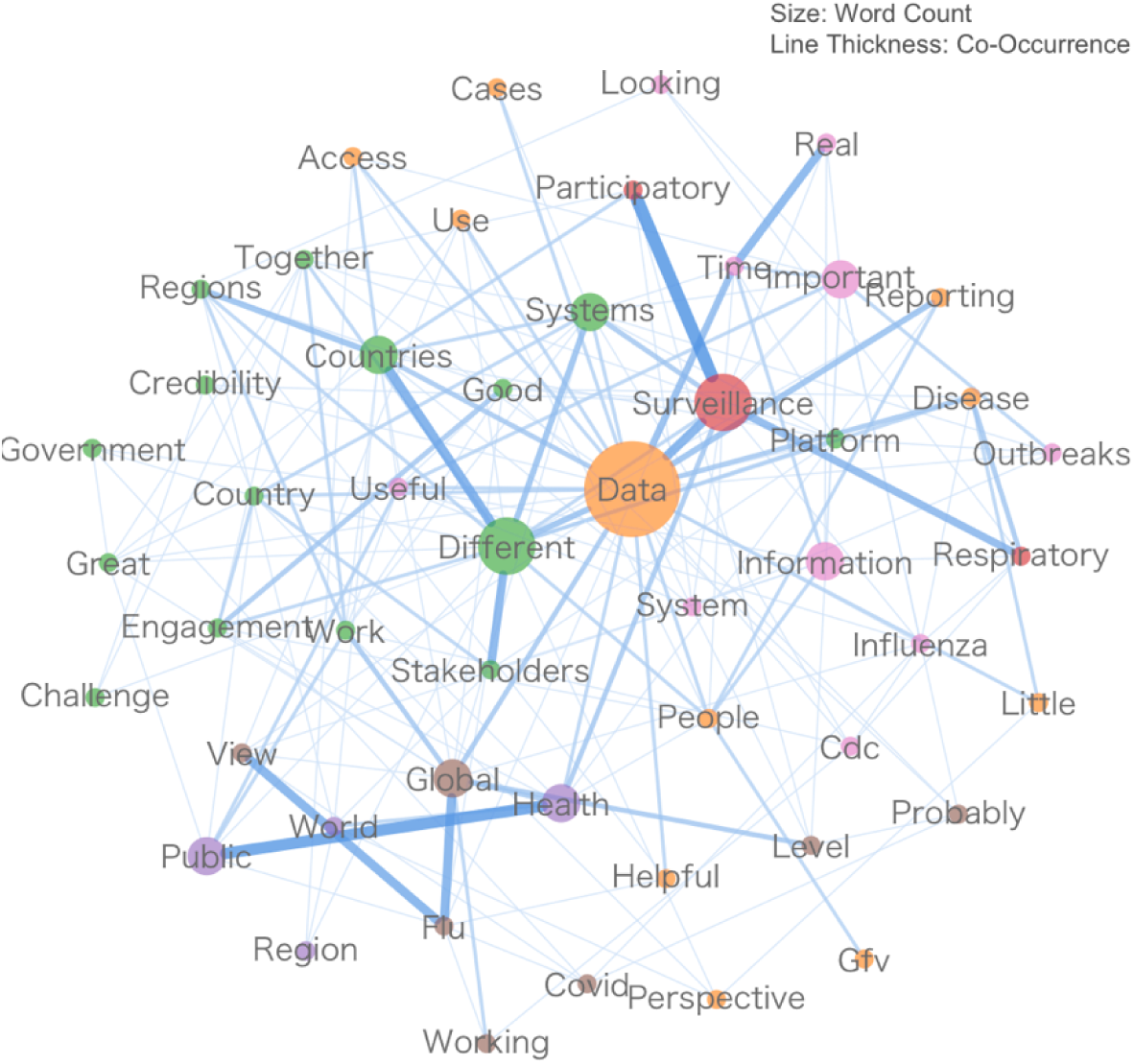
Word pair network from public health researchers

Below are some excerpts from interview transcripts:

> “I think that in principle the API is very interesting and work well, but we face a strong limitation from the European side due to restrictions on data sharing resolution beyond the country level. While countries like Australia and the US are able to share data at the postal code level, allowing users to access more detailed information as they zoom into the map, Europe can only share country-level data. This means we cannot fully utilize the API’s potential, though this is a limitation on our side rather than a technical constraint of Global Flu View.” GFV partner.

> “I think it’s the first platform bringing together data from several different systems across the globe, which has never been done before. The multiple-year process of setting up data sharing agreements to make Global Flu View possible was a huge step forward. I truly believe it is a global orchestrator because it brings together various data sources while allowing people to explore data within individual systems, as well as across multiple combinations of systems or all systems together. With that in mind, I feel it’s a very successful system orchestrator for self-reported influenza-like illness.” Advisory group member.

> “It has the potential to help strengthen our overall global situational awareness of the burden and impact of respiratory disease. Though we’re still some way from achieving this, as it requires substantial capacity building, implementation work, and identification of countries capable of implementing these approaches. Based on my experience, Global Flu View can demonstrate utility and showcase how countries are using this information. It can help advance the field by promoting participatory surveillance as an additional tool in the overall toolbox of respiratory surveillance systems.” Public health researcher

### 3.2. Generative AI sentiment analysis

Sentiment analysis results varied across stakeholder groups. Among GFV partners, ChatGPT generated the highest sentiment score (0.8), followed by Gemini (0.7), Perplexity (0.6), and Claude AI (0.4). For advisory group members, ChatGPT and Gemini both produced scores of 0.8, while Claude AI and Perplexity each rated sentiment at 0.7. Public health researchers’ responses received the highest sentiment score from Gemini (0.8), with ChatGPT rating lower at 0.6, while both Claude AI and Perplexity assessed sentiment at 0.4 (Fig. 5)

**Figure 5.**
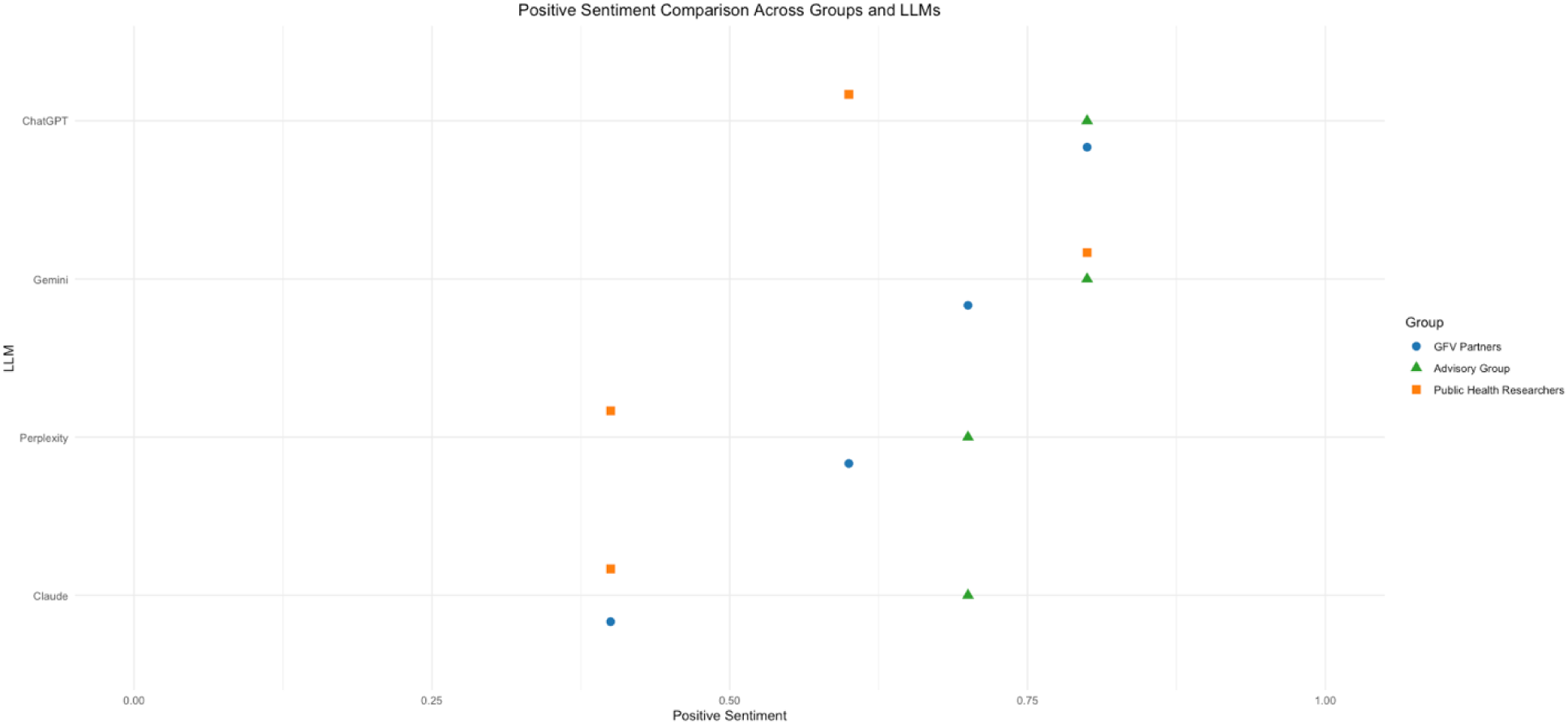
Positive sentiment spectrum according to participants group across LLMs.

### 3.3. Average rank by LLMs

Based on the Friedman ranking (figure 6), ChatGPT and Gemini consistently achieved the top two ranks across all groups, indicating higher sentiment scores overall (average rank = 3.5 each). Claude emerged as the lowest-ranked model (average rank ≈ 1.33), and Perplexity placed slightly higher than Claude (average rank ≈ 1.67). The standard deviations suggest that Claude and Perplexity had relatively stable rankings (SD ≈ 0.29), whereas ChatGPT and Gemini showed moderate variability (SD ≈ 0.50) across the three participant groups.

**Figure 6.**
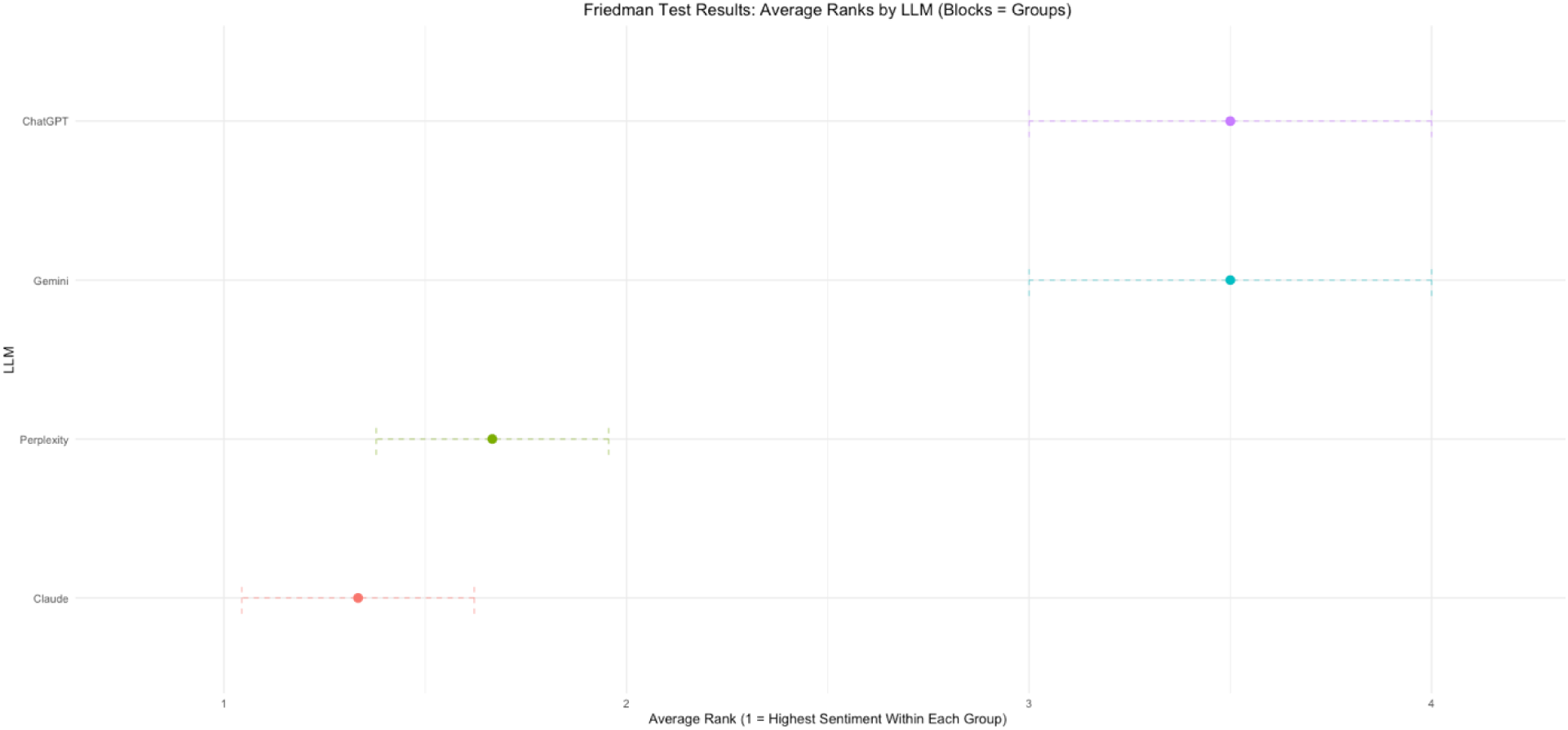
Average ranks by LLM using Friedman test.

## 4. Discussion

The objective of this study was to assess the utility of GFV and gather insights into its perceived effectiveness from the perspective of its partners, advisory group members, and public health researchers, using LLMs. We found that the overall sentiment towards GFV was positive among three stakeholder groups, ranging from 0.4 to 0.8 across the four AI platforms used. Evaluating the utility of GFV from the perspective of persons deemed to be knowledgeable on ILI participatory surveillance systems is paramount to enhance its functionality and reach a greater impact on ILI surveillance. Several studies have been conducted, comparing AI platforms performance in various medical fields [34–38]. However, this study is the first evaluation of GFV run through AI platforms. It utilized four LLMs to run SA on responses from study participants, using a common optimized prompt across all AI platforms. Research has shown that optimized prompts comprising detailed instructions allow ChatGPT to generate enhanced results [32]. Results of SA were presented on a sentiment spectrum ranging from -1 being negative to +1 being positive. Among GFV partners, ChatGPT rated the sentiment of the responses as 0.8, followed by Gemini (0.7), Perplexity (0.6) and Claude AI (0.4). Among advisory group members, both ChatGPT and Gemini rated the sentiment as 0.8, while both Claude AI and Perplexity generated a score of 0.7. Among public health researchers, the score was 0.8 from Gemini, followed by 0.6 from ChatGPT, and 0.4 from both Claude AI and Perplexity. These findings indicate that sentiment varied both across stakeholder groups and AI platforms. ChatGPT and Gemini consistently generated higher sentiment scores across all groups, while Claude AI and Perplexity tended to yield more conservative sentiment scores. The variation in scores between platforms analyzing the same responses suggests that choice of generative AI tools may significantly impact SA results. This underscores the importance of using multiple LLMs for comparison when conducting AI-based SA, since different platforms may interpret emotional content differently. The general positive sentiment scores (> 0.4) across all groups and platforms indicate overall favorable perceptions of GFV, despite notable differences in enthusiasm levels between stakeholder categories.

Sentiment analysis (SA) was used in this study to evaluate opinions of participants on GFV. SA is the computational study of persons’ opinions, attitudes and emotions toward an entity [39]. Research has shown ChatGPT demonstrating incredible natural language understanding, generation and capabilities of solving complex reasoning tasks [40]. There are two mechanisms allowing ChatGPT to contribute significantly to SA. First, its advanced semantic comprehension, logical inference comprehension and logical inference capabilities allow it to interpret contextual meanings and determine the underlying reasons for specific sentiments, which enhances analysis accuracy. Additionally, since ChatGPT is built on LLM architecture, it can be adapted through fine-tuning on extensive sentiment-labeled datasets to recognize subtle patterns in how emotions and opinions are expressed [41].

The practical implications of our findings are twofold, impacting both the GFV platform and the methodology employed in SA. At GFV level, the consistently positive sentiment scores across stakeholder groups signify strong approval and trust in the platform’s capability to enhance ILI surveillance. The nuanced variations in sentiment between stakeholder groups can guide targeted platform improvements, indicating potential opportunities for enhancement. At the methodological level, our approach of using multiple LLMs for SA reveals important considerations for future AI-assisted qualitative research. The systematic variation in sentiment scores between platforms demonstrates that the selection of platforms can impact research outcomes. This finding reinforces the importance of using multiple AI platforms for comparison in SA, like how traditional qualitative research often uses multiple coders to ensure reliability. Furthermore, our use of a standardized prompt across different LLMs provides a methodological framework for future studies looking to use AI for qualitative analysis while keeping consistency and comparability.

This comprehensive analysis using multiple AI platforms and SA has provided valuable insights while also demonstrating the potential of LLMs in public health research evaluation. As both surveillance platforms and AI capabilities continue to advance, the methodology employed in this study could serve as a model for future evaluations of public health tools and platforms. The Friedman ranking revealed discernible differences in sentiment scores among the four LLMs, as evidenced by distinct average rank values. In particular, Claude consistently achieved the lowest average rank, indicating relatively lower sentiment levels across the three user groups. By contrast, Gemini and ChatGPT both occupied the highest average ranks, suggesting that they produced higher sentiment outputs more consistently. Perplexity fell between these two extremes, with moderately higher sentiment than Claude but still lower than Gemini and ChatGPT. Notably, the standard deviations of these ranks indicate that Claude’s and Perplexity’s positions were more stable across groups, while Gemini and ChatGPT showed slightly more variability. Overall, these findings imply that certain LLMs may be better suited to tasks or contexts requiring higher expressed sentiment, whereas others may have a tendency toward more neutral or lower sentiment output. Further investigation with larger sample sizes or additional benchmarking tasks would help validate these trends and explore the possible drivers behind each model’s sentiment behavior.

This study had some limitations. First, the small sample size of 10 participants, despite inviting 17 stakeholders, may not provide a comprehensive representation of all perspectives on the GFV platform. The limited number of participants in each stakeholder category (5 GFV partners, 2 advisory members, and 3 researchers) particularly affects the generalizability of subgroup analyses. Second, our approach to SA using LLMs had inherent limitations. We employed a single optimized prompt across all four platforms, which may not have captured the full complexity of participants’ sentiments. The lack of validation against human coders and absence of inter-model reliability assessment limits our ability to verify the accuracy of the SA results. Furthermore, the inherent biases and limitations of LLMs in interpreting nuanced human responses must be acknowledged. Finally, the study may have provided only a snapshot view of GFV’s utility. As the platform will continue to evolve and user needs will change, perspectives captured in this study may only reflect part of current sentiments.

Looking forward, our findings suggest several promising directions for both GFV development and research methodology. Future studies should investigate the factors driving these variations between platforms and establish best practices for standardizing AI-based SA in qualitative research. Additionally, assessing the impact of different prompt engineering approaches on AI-based sentiment analysis could enhance the methodology for future evaluations. As participatory surveillance continues to evolve in the global health landscape, the integration of AI-based evaluation methods with traditional qualitative analysis may provide increasingly robust frameworks for platform assessment.

## 5. Conclusion

In summary, this study is the first evaluation of GFV utilizing LLMs for SA, revealing generally positive and optimistic perceptions of the platform’s utility. The analysis across four LLMs (ChatGPT, Claude AI, Perplexity, and Gemini) showed a general positive sentiment score towards GFV, with some variations across stakeholder groups. These scores indicate that there is room for enhancement of the platform. Despite the study’s limitations, these results provide valuable insights for the future development of GFV, suggesting that with continued refinement and attention to stakeholder feedback, GFV can strengthen its role in global influenza surveillance and public health decision-making.

## Acknowledgement

We would like to thank Ending pandemics and the Skoll foundation for funding this study.

## Author contributions

**Conceptualization:** JL, OLN

**Data collection**: JL

**Data curation**: JL

**Data analysis:** JL, OLN

**Funding acquisition:** OLN, IH

**Writing - original draft:** JL

**Writing – review & editing:** OLN, IH

## AI Statement

AI tools were used for improving the language and proofreading the text.

## Data availability statement

De-identified datasets may be provided upon request to the corresponding author.

